# Epigenome-wide association study, meta-analysis and risk profiling of whole blood in Parkinson’s disease

**DOI:** 10.1101/2024.05.28.24308034

**Authors:** Ingeborg Haugesag Lie, Manuela Tan, Maren Stolp Andersen, Mathias Toft, Lasse Pihlstrøm

## Abstract

An increasing body of evidence indicates altered DNA methylation in Parkinson’s disease (PD), yet the reproducibility and utility of such methylation changes are largely unexplored. We conducted an epigenome-wide association study (EWAS) in whole blood, including 280 PD and 279 control participants from Oslo, Norway. In meta-analysis with data from the Parkinson’s Progression Markers Initiative (PPMI) and a previously published whole blood PD EWAS (total N=3068) we confirm *SLC7A11* hypermethylation and nominate a novel suggestive differentially methylated CpG near *LPIN1*. A joint multiscore risk profiling model incorporating polygenic risk and methylation-based estimates of epigenetic PD risk, smoking and leukocyte proportions differentiated patients from control participants with an area under the receiver-operator curve or 0.82 in the Oslo cohort and 0.65 in PPMI. Our results highlight the power of DNA methylation profiling to capture multiple aspects of disease risk, indicating a biomarker potential for precision medicine in neurodegenerative disorders.

## Introduction

Parkinson’s disease (PD) is a progressive neurodegenerative disorder with no available disease modifying treatment (1). It is the most common movement disorder and second most common neurodegenerative disease globally, affecting over 8.5 million people with a prevalence between 1-2% in the population above 65 years of age (2-4). The prevalence has more than doubled since 1990, mainly driven by aging populations and longer life expectancy (3, 5, 6). It is estimated that the number of affected individuals will continue to increase (3, 5). There is currently an urgent need for an improved understanding of the molecular disease mechanisms and novel biomarkers to stratify patients and track disease progression.

DNA methylation at CpG dinucleotides is an important mechanism of gene regulation and currently widely studied in the context of complex disease using a hypothesis-free epigenome-wide association study (EWAS) methodology (7). However, methylation is cell-type specific, dynamic over time and shaped by both genetic and environmental factors, complicating EWAS design and interpretation (7, 8). In recent years, a number of EWAS have reported differential methylation in brain tissue or isolated neurons from PD donors (9-12), yet the availability of neuronal tissue is limited. In contrast, whole blood is readily accessible and arguably more relevant from a biomarker perspective. Several whole blood EWAS have been reported in PD (13-16), the largest including more than 2000 participants (13).

The increasing availability of epigenome-wide methylation data holds promise for risk stratification and individualized genomic profiling, which could improve PD care within the framework of precision medicine. Estimating the cumulative burden across many independently disease-associated single-nucleotide polymorphisms (SNPs) as a polygenic risk score (PRS) is currently an established standard methodology in complex disease genetics (17). Similarly, Vallerga et al. showed that methylation-based risk scores modelled on one whole blood EWAS was associated with PD status when tested as an out-of-sample classifier in an independent EWAS dataset (13). Furthermore, methylation-based scoring algorithms have been used to estimate accelerated epigenetic aging and cell-type proportions across classes of leukocytes, both showing association with PD (13, 18). However, the prospect of integrating multiple methylation-based scores for risk prediction has not been systematically explored in PD.

The objectives of this study were to further elucidate the role of dysregulated DNA methylation in PD and to evaluate the potential of methylation profiling for predicting disease status. We conducted a whole blood EWAS of 280 PD participants and 279 healthy controls from Oslo, Norway, using the Illumina MethylationEPIC array (19). We also analyzed publicly available complementary case-control data from the Parkinson’s Progression Markers Initiative (PPMI) study (20) and compared and meta-analyzed results with the meta-EWAS previously published by Vallerga et al (13), thus to the best of our knowledge incoporating all available large datasets of whole blood methylation in PD. We show that although the reproducibility of individual top-probe associations seems limited across studies, different scoring algorithms capture unique aspects of disease risk that can be integrated in a powerful combined epigenetic and genetic risk score.

## Results

### EWAS analysis of differentially methylated CpGs in the Oslo cohort

We recruited 280 individuals with sporadic PD and 279 controls without neurological disease at Oslo University Hospital, Norway. All were unrelated and of European descent. Notably, the PD participants had a relatively early mean age at onset (52.9 years) and a mean disease duration of 10.3 years at the time of blood sampling. Further demographic details are provided in Table 1.

**Table 1:** Cohort demographics. **Abbreviations:** SD = Standard deviation, PD = Parkinson’s disease, PPMI = Parkinson’s Progression Markers Initiative

| Status |  | Female | Age at visit |  |  | Age at onset |  |  | Disease duration |  |  |
| --- | --- | --- | --- | --- | --- | --- | --- | --- | --- | --- | --- |
|  | N (%) | N (%) | Mean | Range | SD | Mean | Range | SD | Mean | Range | SD |
| **Oslo (n=559)** |  |  |  |  |  |  |  |  |  |  |  |
| PD | 280<br>(50.0) | 91<br>(0.33) | 63.2 | 35 - 83 | 9.1 | 52.9 | 24 - 81 | 10.1 | 10.3 | 0 - 39 | 6.4 |
| Controls | 279<br>(50.0) | 80<br>(0.29) | 61.8 | 40 - 91 | 11.3 |  |  |  |  |  |  |
| All | 559<br>(100) | 171<br>(0.31) | 62.5 | 35 - 91 | 10.3 |  |  |  |  |  |  |
| **PPMI (n=378)** |  |  |  |  |  |  |  |  |  |  |  |
| PD | 269<br>(0.71) | 92<br>(0.34) | 61.9 | 37 - 83 | 9.3 | 59.9 | 35 - 81 | 9.5 | 1.9 | 0 - 20 | 1.9 |
| Controls | 109<br>(0.29) | 29<br>(0.27) | 62.1 | 31 - 83 | 10.6 |  |  |  |  |  |  |
| All | 378<br>(100) | 121<br>(0.32) | 62.0 | 31 - 83 | 9.7 |  |  |  |  |  |  |

Epigenome-wide methylation was analyzed using the Illumina MethylationEPIC array (19). Following quality checks, filtering and normalization (see Methods) our final dataset included 497,578 methylation beta values in 559 individuals. Statistical hypothesis testing was performed using mixed linear model-based omic association (MOA) as implemented in the software package OSCA (21), adjusting for sex, age, estimated cell composition and experiment plate (see Methods). The same MOA approach was previously applied in the largest whole blood EWAS of PD to date (13) and aims to eliminate potential unobserved confounding and control genomic inflation by adjusting for the random effect of total genome-wide DNA methylation (13, 21). We observed 13 probes passing the recommended epigenome-wide significance threshold of p<9*10^-8^ (22) in the Oslo dataset. (Figure 1, Table 2). The genomic inflation factor was λ=1.009.

**Figure 1.**
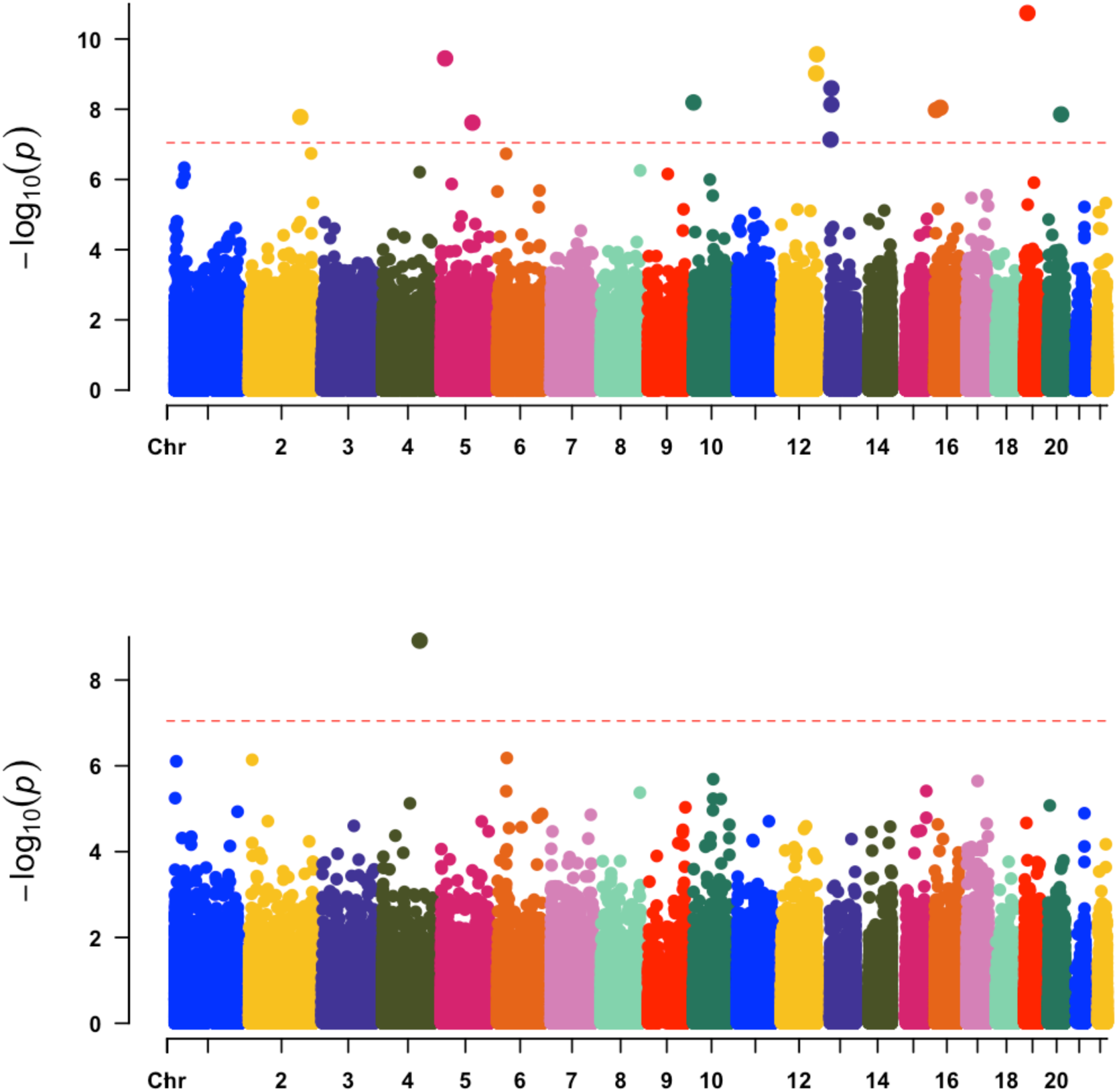
Manhattan plots. Manhattan plots of p-values from mixed linear model-based omic association analysis (MOA) in the Oslo cohort (top) and fixed effect meta-analysis of Oslo, Parkinson’s Disease Progression Marker Initiative (PPMI) and Vallerga et al. summary statistics (bottom). The dashed line indicates the recommended Bonferroni-adjusted threshold for MethylationEPIC array experiments at p<9*10^-8^.

**Table 2.**
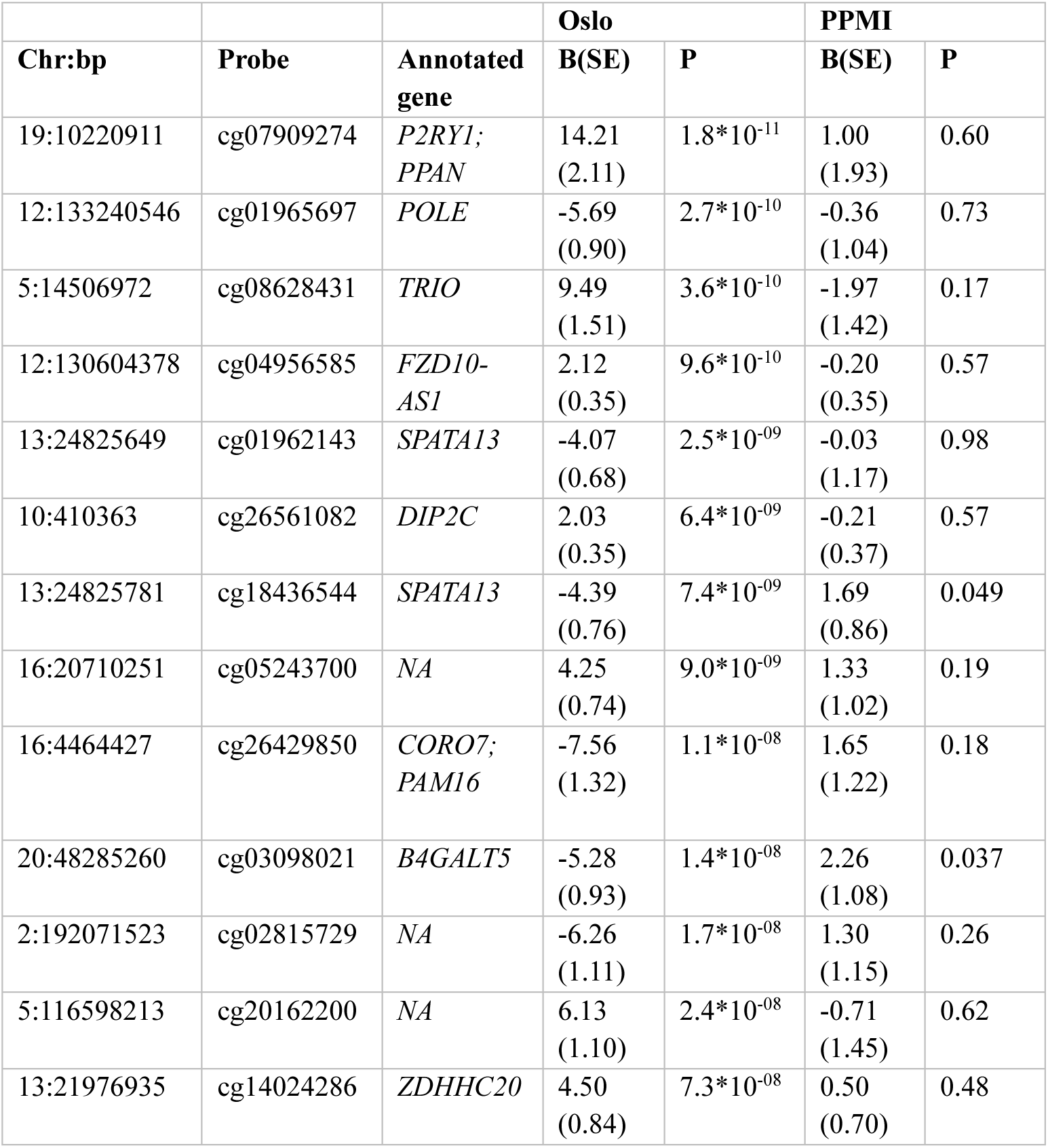
CpG-probes passing the p<9*10^-8^ significance threshold in the Oslo cohort. The table shows results from mixed linear model-based omic association analysis (MOA) performed using the software package OSCA in the Oslo and PPMI cohorts. **Abbreviations:** PPMI = Parkinson’s Progression Marker Initiative, B = Regression coefficient, P = P-value, SE = Standard error

### No replication of single-probe associations in the PPMI dataset

Aiming to compare association results and potentially replicate findings in an independent case-control dataset, we downloaded raw intensity files from an Illumina MethylationEPIC experiment performed in DNA from whole blood including 269 PD participants and 109 controls from PPMI, as well as clinical and genetic data from the same individuals. We note that sampling for this experiment was performed at multiple centers, on average 1.9 years after disease onset in the PD participants, compared to 10.3 years in Oslo. Mean age at onset was 7 years higher in PPMI than in the Oslo cohort, indicating major differences in cohort characteristics across these two studies (Table 1). We processed the methylation data using an identical bioinformatic pipeline as the Oslo dataset (see Methods). MOA analysis was performed adjusting for sex, age and estimated cell proportions. None of the 13 probes nominated in the Oslo dataset were replicated in PPMI data (Table 2) (λ=0.91). No probe reached the epigenome-wide significance level in the PPMI data alone (λ=0.91). We then performed meta-analysis of the Oslo and PPMI results, but observed no associations passing the significance threshold.

### SLC7A11 association replicates in Oslo data, but not in PPMI

Next, we compared our results to summary statistics from the large whole blood PD EWAS published by Vallerga et al., which included 1,132 PD participants and 999 controls from two independent cohorts, with disease duration more similar to the Oslo dataset (13). This study was based on the Illumina Infinium Human Methylation 450K BeadChip, which contains fewer CpG probes than the more recent 850K MethylationEPIC array. The published summary statistics includes data on 229,071 probes. Only one out of the 13 probes nominated in the Oslo dataset was present in the Vallerga summary statistics, showing no association with PD (p=0.23 for cg04956585).

Vallerga et al. reported a significant hypermethylation signal from cg06690548 in the *SLC7A11* locus (13). Consistent with this result, we found evidence of cg06690548 hypermethylation in Oslo PD participants (coeff(SE)=2.37(0.48), p=6.2*10^-07^). However, no association was seen for the same probe in PPMI data (coeff(SE) = -0.28(0.42), p=0.50).

To assess the degree of consistency in differential methylation also for probes associated at sub-significant p-values, we generated beta-beta-plots comparing the coefficients of the 50 most significant probes from the Vallerga et al. summary statistics against their coefficients in the Oslo and PPMI analyses respectively (Figure 2). In the Oslo dataset, 36 out of 50 probes showed a direction of effect consistent with the Vallerga et al study and coefficients showed a clear correlation (r^2^=0.54, p=4.3*10^-5^). In contrast, PPMI data showed no trend towards consistent directions of effect nor correlated probe coefficients (r^2^ =-0.21, p=0.15).

**Figure 2.**
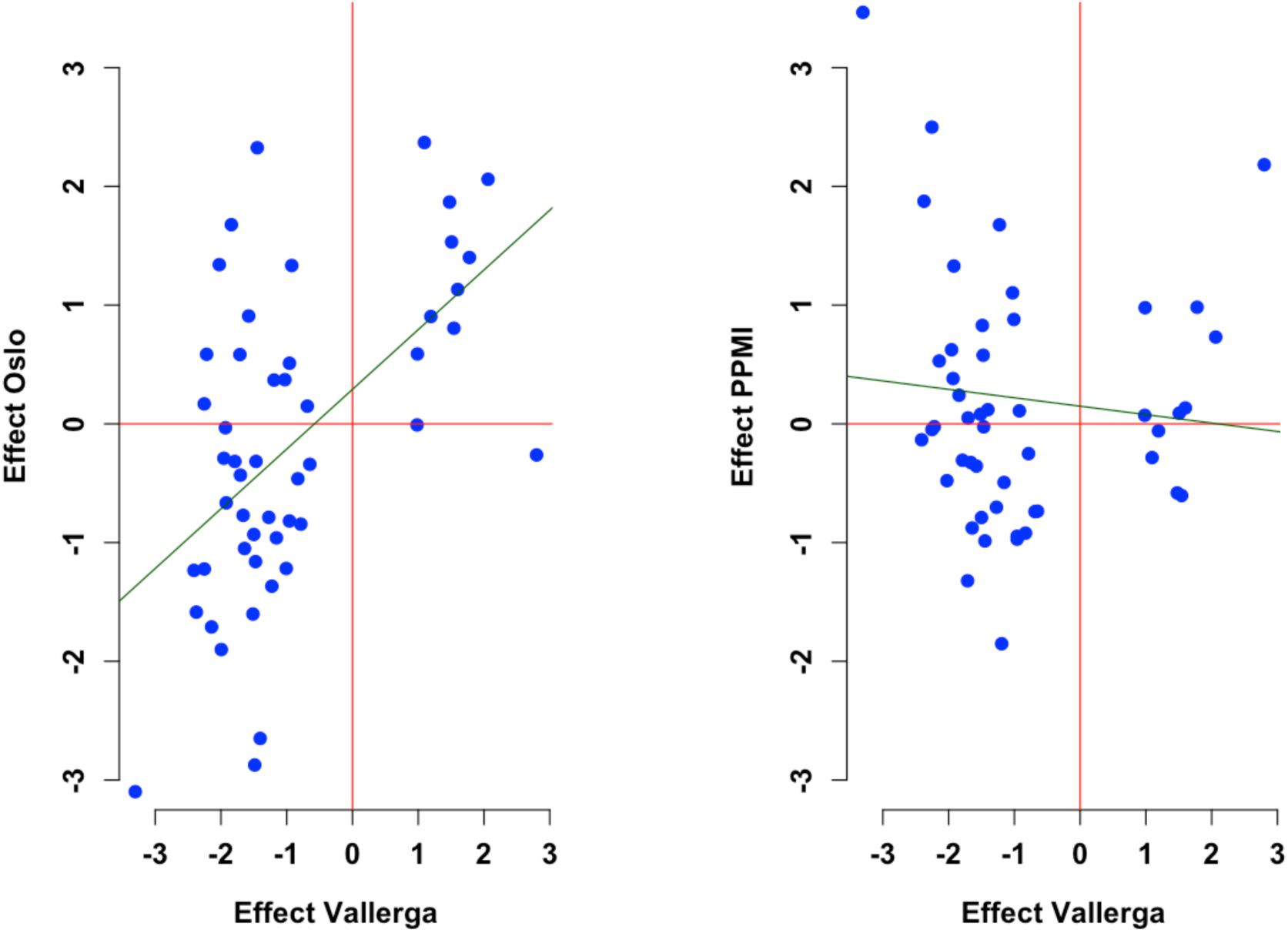
Beta-beta plots comparing effect estimates from Vallerga et al. to Oslo and PPMI results. Out of the CpG probes that overlap between all three datasets, we selected the 50 probes with the lowest p-values in the previously published summary statistics from Vallerga et al. and plotted reported effect estimates from this study against corresponding estimates in Oslo and Parkinson’s Progression Marker Initiative (PPMI) data. The results show a clear correlation of effects between Vallerga et al. and Oslo data (r2 = 0.54, p=4.3*10^-5^), but not between Vallerga et al. and PPMI (r2 = -0.21, p=0.15).

### Meta-analysis nominates a novel suggestive association near LPIN1

We meta-analyzed results from all three studies, including a total of 148,579 probes present in all datasets (λ=1.02). The previously highlighted *SLC7A11* locus (cg06690548) was the only signal to reach the suggested significance threshold for the MehylationEPIC array (p<9*10^-8^) in this meta-analysis, driven by the association in the Vallerga and Oslo datasets (coeff(SE)=1.04(0.17), p=1.2*10^-09^) (Table 3). Given that the number of probes tested for association in this meta-analysis was considerably smaller, we also explored a more liberal approach to multiple testing adjustment. Applying Benjamini-Hochberg correction (23), we found 4 CpG probes with a false discovery rate (FDR)<0.05 (Table 3). One of these, cg10142874 located in an intron of *LPIN1*, was the only probe to reach nominal significance (p<0.05) in all three cohorts, indicating possible hypermethylation near the time of diagnosis (PPMI baseline visit) (Table 3).

**Table 3.** CpG-probes passing the FDR-corrected threshold in meta-analysis. The table shows results from mixed linear model-based omic association analysis (MOA) performed using the software package OSCA. Fixed effect meta-analysis was performed for 148,579 probes overlapping in the Oslo, PPMI and Vallerga summary statistics. **Abbreviations:** PPMI = Parkinson’s Progression Marker Initiative, B = Regression coefficient, P = P-value, SE = Standard error, FDR = Benjamini-Hochberg False discovery rate

| Chr:bp | Probe | Annotated gene | Oslo |  | PPMI |  | Vallerga |  | Meta |  |  |
| --- | --- | --- | --- | --- | --- | --- | --- | --- | --- | --- | --- |
|  |  |  | B(SE) | P | B(SE) | P | B(SE) | P | B(SE) | P | FDR |
| 4:139162808 | cg06690548 | *SLC7A11* | 2.37<br>(0.48) | $6.2 \times 10^{-07}$ | -0.28<br>(0.42) | 0.5<br>0 | 1.09<br>(0.20) | $6.2 \times 10^{-08}$ | 1.04<br>(0.17) | $1.2 \times 10^{-09}$ | 0.00018 |
| 6:35654363 | cg03546163 | *FKBP5* | -0.34<br>(0.36) | $3.4 \times 10^{-01}$ | -0.74<br>(0.46) | 0.11 | -0.65<br>(0.14) | $2.7 \times 10^{-06}$ | -0.62<br>(0.12) | $6.6 \times 10^{-07}$ | 0.029 |
| 2:11917623 | cg10142874 | *LPIN1* | 2.94<br>(0.85) | $5.4 \times 10^{-04}$ | 1.94<br>(0.82) | 0.0<br>18 | 1.40<br>(0.45) | $2.0 \times 10^{-03}$ | 1.78<br>(0.36) | $7.2 \times 10^{-07}$ | 0.029 |
| 1:6341230 | cg21220721 | *ACOT7* | 1.44<br>(0.45) | $1.4 \times 10^{-03}$ | 0.96<br>(0.55) | 0.0<br>79 | 0.86<br>(0.24) | $4.2 \times 10^{-04}$ | 0.98<br>(0.20) | $7.8 \times 10^{-07}$ | 0.029 |

### Evaluating a methylation-based PD risk score

Having observed some degree of consistency across the Vallerga and Oslo datasets with respect to differential methylation of individual probes, we next assessed a number of scoring algorithms that take advantage of broader sets of methylation values weighted against different types of reference data for methylation-based profiling. We used the *score* function in OSCA (21) with the summary statistics from the meta-analysis published by Vallerga et al.(13) as weights to generate individual methylation-based risk scores, also known as methylation profile scores (MPS), for all participants in the Norwegian cohort. The z-normalized MPS was significantly associated with PD case/control-status, with odds ratio (OR) 1.29, 95% confidence interval (CI) (1.09-1.54), p=0.0042, controlled for age and sex. The area under the receiver-operator curve (AUC) was 0.59 (95% CI =0.54-0.64) (Table 4). We note however that the MPS had lower performance as an out-of-sample classifier in our data compared to the Vallerga et al. study, where the reported AUC was 0.70 (95% CI =0.66-0.75) (13).

**Table 4.** Association of individual methylation-based and polygenic scores with Parkinson’s disease. Results from logistic regression analysis and estimates of area under the receiver operator curve. Scores were Znormalized to the mean value in controls. Odds ratios and P-values are reported for logistic regression models including age and sex as covariates. **\*** In the PPMI-cohort, the logistic regression models on DNA methylation-scores are performed on n=378 individuals, the model including PRS is performed on n=342 with available genetic data. **Abbreviations:** P=P value, AUC= Area under the receiver operator curve, OR = Odds ratio, 95% CI = Confidence interval

|  |  | Oslo (N=559) |  | PPMI (N=378*) |  |  |
| --- | --- | --- | --- | --- | --- | --- |
| Score | OR<br>(95%CI) | P | AUC,<br>(95% CI) | OR<br>(95%CI) | P | AUC<br>(95% CI) |
| Polygenic risk score | 1.70<br>(1.42 - 2.04) | 9.9*10<sup>-09</sup> | 0.64<br>(0.60 - 0.69) | 1.65<br>(1.31 - 2.10) | 2.8*10<sup>-05</sup> | 0.65<br>(0.58 - 0.71) |
| Methylation Profile Score | 1.29<br>(1.09 - 1.54) | 0.0042 | 0.59<br>(0.54 - 0.64) | 1.01<br>(0.76 - 1.37) | 0.95 | 0.49<br>(0.42 - 0.56) |
| Smoking Score | 0.51<br>( 0.41 - 0.63) | 1.2*10<sup>-09</sup> | 0.65<br>(0.61 - 0.70) | 0.81<br>( 0.63 - 1.03) | 0.085 | 0.54<br>(0.48 - 0.60) |
| Epigenetic Age acceleration | 1.00<br>(0.83 - 1.20) | 0.99 | 0.50<br>(0.45 - 0.55) | 0.90<br>(0.71 - 1.14) | 0.38 | 0.48<br>(0.42 - 0.54) |
| Granulocyte proportion | 3.46<br>(2.66 - 4.58) | < 2*10<sup>-16</sup> | 0.74<br>(0.70 - 0.78) | 1.43<br>(1.14 - 1.79) | 0.0019 | 0.60<br>(0.54 - 0.66) |
| CD4+ T-cell proportion | 0.46<br>(0.36 - 0.57) | 3.8*10<sup>-12</sup> | 0.67<br>(0.63 - 0.72) | 0.80<br>(0.65 - 0.98) | 0.034 | 0.57<br>(0.51 - 0.63) |
| CD8+ T cell proportion | 0.68<br>(0.55 - 0.83) | 0.00020 | 0.57<br>(0.52 - 0.62) | 0.84<br>(0.67 - 1.06) | 0.14 | 0.46<br>(0.40 - 0.53) |
| Monocyte Proportion | 0.39<br>(0.30 - 0.50) | 3.6*10<sup>-13</sup> | 0.68<br>(0.64 - 0.72) | 0.86<br>(0.69 - 1.09) | 0.21 | 0.55<br>(0.49 - 0.62) |
| NK cell Proportion | 0.66<br>(0.53 - 0.81) | 7.4*10<sup>-05</sup> | 0.58<br>(0.53 - 0.63) | 1.07<br>(0.85 - 1.36) | 0.56 | 0.52<br>(0.46 - 0.58) |
| B cell Proportion | 0.50<br>(0.40 - 0.62) | 1.9*10<sup>-09</sup> | 0.65<br>(0.61 - 0.70) | 0.55<br>(0.33 - 0.86) | 0.017 | 0.59<br>(0.53 -0.65) |

### Association between leukocyte cell type proportions and PD

Several studies have consistently demonstrated that PD patients have higher levels of granulocytes compared to healthy controls (13, 15, 18). We estimated cell type proportions (CTPs) in whole blood using the reference data and algorithm from Houseman et al. as implemented in the *minfi* R package (24, 25). In line with previous reports (13, 18), we found that PD subjects had a significantly higher proportion of granulocytes and lower proportions of B-cells, helper (CD4+) T cells and cytotoxic (CD8+) T cells compared to controls (Extended Data Figure 1, Extended Data Table 1). Interestingly, we also found the estimated proportion of monocytes and natural killer cells (NK-cells) to be significantly lower among the PD subjects. The six estimated CTPs were individually associated with PD case/control-status in logistic regression analyses, correcting for age and sex (Table 4). Pairwise correlation analyses showed that many of the CTPs were linearly correlated, which is expected as these are estimates of proportions that equal a total of 1.0, rather than absolute counts (Extended Data Figure 2).

### Estimating accelerated epigenetic aging

A subset of CpGs have been shown to be associated with chronological age, forming the basis of an “epigenetic clock” (26). The epigenetic clock algorithm estimates epigenetic age, taking DNA methylation data from blood or other tissues as input (26). Previous studies have demonstrated an association between accelerated epigenetic age in whole blood and PD (18), yet this finding has not been consistently replicated (27). We estimated epigenetic age using Horvath’s clock and calculated the measure for age acceleration as described in previous studies (26-28). We found no significant association between epigenetic age acceleration and PD case/control-status in the Oslo dataset, corrected for age and sex (coef =-0.00031, p=0.99).

### A methylation-based smoking score

Tobacco smoking has consistently shown an inverse association with PD across epidemiological studies (29). As smoking has a distinct footprint on DNA-methylation, the likely impact of smoking on an individual’s epigenome can be estimated from methylation data (30-32). We used the R package EpiSmoker (32) to generate DNA methylation smoking scores (SSc) based on the algorithm by Elliot et al. (31) for all study participants. The SSc is a continuous score associated with smoking and the higher the score, the higher the probability for current smoking. In line with the well-established epidemiological observations, we found the SSc to be negatively associated with PD status in the Oslo cohort (coef=-0.15, p=1.2*10^-09^) adjusted for sex and age (Table 4). Data on self-reported smoking was available only for a minor subset of subjects (N PD/Controls= 24/24). Only one of these participants (control) reported current smoking, and very few subjects reported previous regular smoking (N PD/Controls=1/2), so we did not have sufficient data to validate the accuracy of the smoking score algorithm in our sample. Interestingly, the case-control difference in SSc was similar for this small group of self-reported never-smokers (N PD/controls=23/22) as in the full Oslo data, although the difference was non-significant with only 45 individuals (p=0.316) (Figure 3). This might raise the hypothesis that the epigenetic smoking score algorithm captures elements of a methylation profile that may be associated with differential PD risk also via other mechanisms than through smoking exposure itself. Analysis of the PPMI cohort showed similar trends between PD and controls, although the differences between groups were not significant (p=0.084 for the whole cohort (N=378) and p=0.21 for the never-smokers (N=112)) (Extended Data Figure 3).

**Figure 3.**
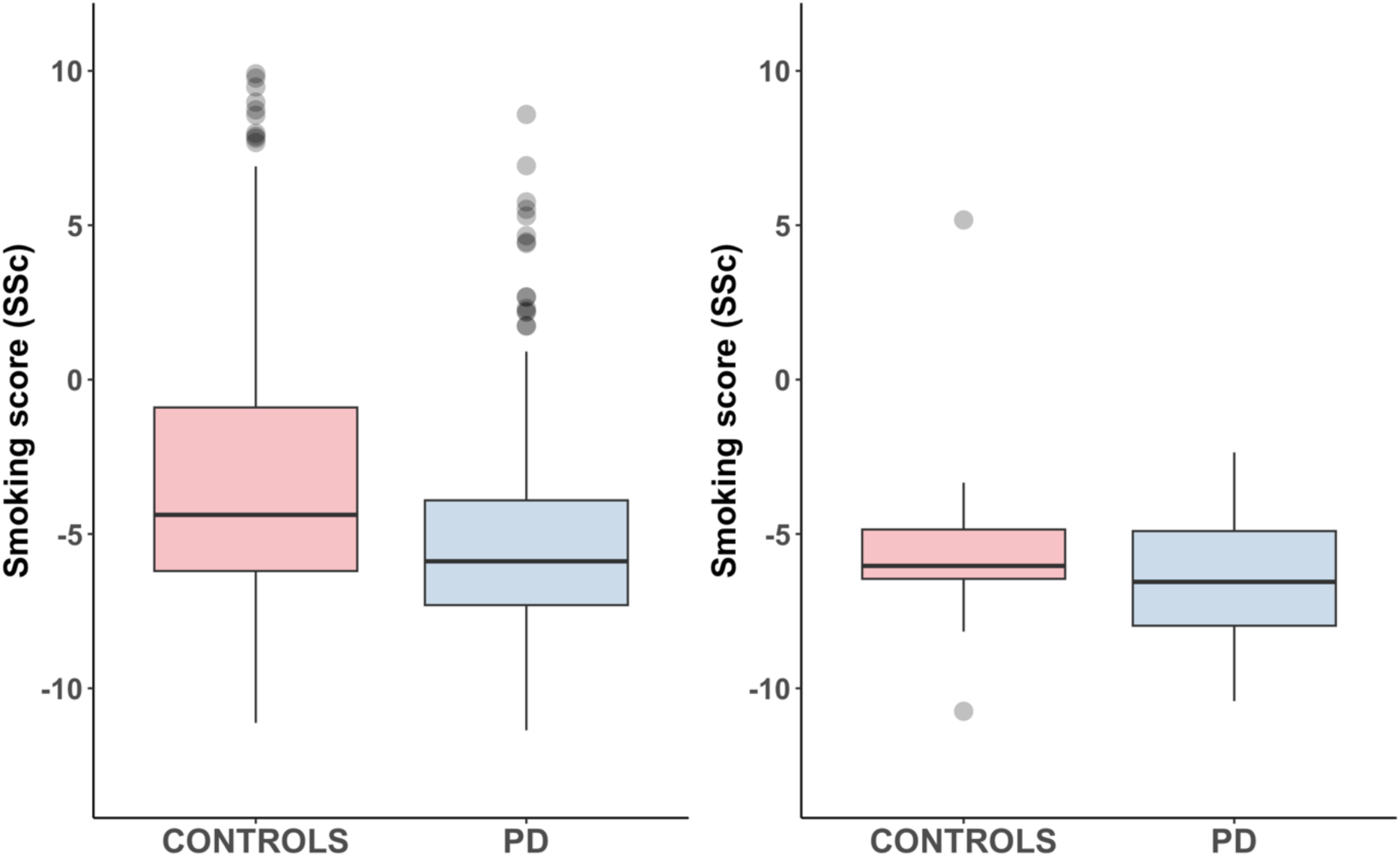
Boxplots comparing Smoking Score between PD and control participants. The boxplots show Smoking Score estimated from whole blood methylation values using the R package EpiSmokEr in all Oslo participants (left, N=559) and self-reported non-smokers only (right, N=45). In logistic regression analysis adjusting for sex and age, the groupwise difference was highly significant in the full dataset (p=1.2*10^-9^), but not for the small set of known self-reported non-smokers (p=0.316).

### Different disease-associated methylation profiles and polygenic burden have additive effects on PD risk

Having identified multiple methylation-based measures that are significantly associated with disease status, we next aimed to assess to what extent these scores capture overlapping or unique aspects of disease risk, and how they relate to the cumulative genetic risk conferred by common susceptibility alleles identified in GWAS. Scatterplots and coefficients from pairwise correlation analysis of all scores are shown in Extended Data Figure 2. Of note, several strong pairwise correlations were seen for the different classes of leukocytes. This was expected, as the algorithm per definition provides estimates as a proportion of all cells in whole blood rather than absolute levels. The different classes of lymphocytes (B cells, CD8+, CD4+ and NK cells) all had Pearson’s R^2^ < -0.5 with granulocytes, whereas monocytes showed a more moderate correlation with granulocytes (Pearson’s R^2^ = -0.32).

The methylation-based PD-risk score (MPS) was not correlated with the polygenic PD-risk score (PRS) (Pearson’s R^2^ = 0.04, p=0.35), indicating that the differential methylation captured in whole blood PD EWAS is not generally driven by the effects of common risk SNPs on DNA methylation (Extended Data Figure 3). Granulocyte-, monocyte- and B-cell proportions were all significantly correlated with PRS in the expected direction in the whole dataset, but not when patients and controls were analyzed individually. The MPS was not correlated to cell type proportions, but significantly positively correlated with the SSc (Pearson’s R^2^=0.256, p=8.3*10^-10^), which was the only correlation we observed with a paradoxical direction relative to the effect on PD risk (Extended Data Figure 2).

After observing several significant pairwise correlations between scores, we assessed their association with PD in a joint logistic regression model, excluding lymphocyte proportions to avoid collinearity. MPS, granulocyte proportion, monocyte proportion, SSc and PRS all remained individually significant in a model adjusting for sex and age (Extended Data Table 2).

### Evaluating combined polygenic-epigenetic profiling for risk prediction

Our findings indicate that from genotypes and methylation array data, multiple profiles can be calculated that carry independent power to differentiate PD patients from controls. To further explore the potential of risk profiling integrating multiple scores, we evaluated the performance of multivariate logistic models for out-of-sample classification in the Oslo cohort. The weights used to estimate MPS and PRS were based on summary statistics from previously published EWAS (13) and GWAS (33), respectively. To reduce the risk of overfitting when modeling the weight of each included variable in a multiscore risk profile, we randomly assigned PD patients and controls to a Training and Test subset (proportioned 0.8/0.2) over 1000 iterations. The coefficients from logistic regression adjusted for sex and age in the Training subset were used to estimate a risk score in the Test set. The performance of the risk score in assigning participants to the PD or control group in the Test set was assessed by calculating the area under the receiver-operator curve (AUC).

We observed increasing performance as more scores were incorporated into the joint risk profile (Extended Data Table 3). The combination of PRS and MPS has been assessed in previous studies of other complex traits with variable results (8, 34-36). We found that a risk profile integrating both MPS and PRS outperforms each of these scores alone, although the added value of MPS was modest. A marked stepwise increase in performance was further observed as Ssc, granulocyte proportion and monocyte proportion were successively included in the model (Extended Data Table 3, Figure 4). The risk profile incorporating all PD-associated scores (PRS, MPS, SSc, granulocyte proportion and monocyte proportion) showed the best performance, with a mean AUC of 0.82.

**Figure 4.**
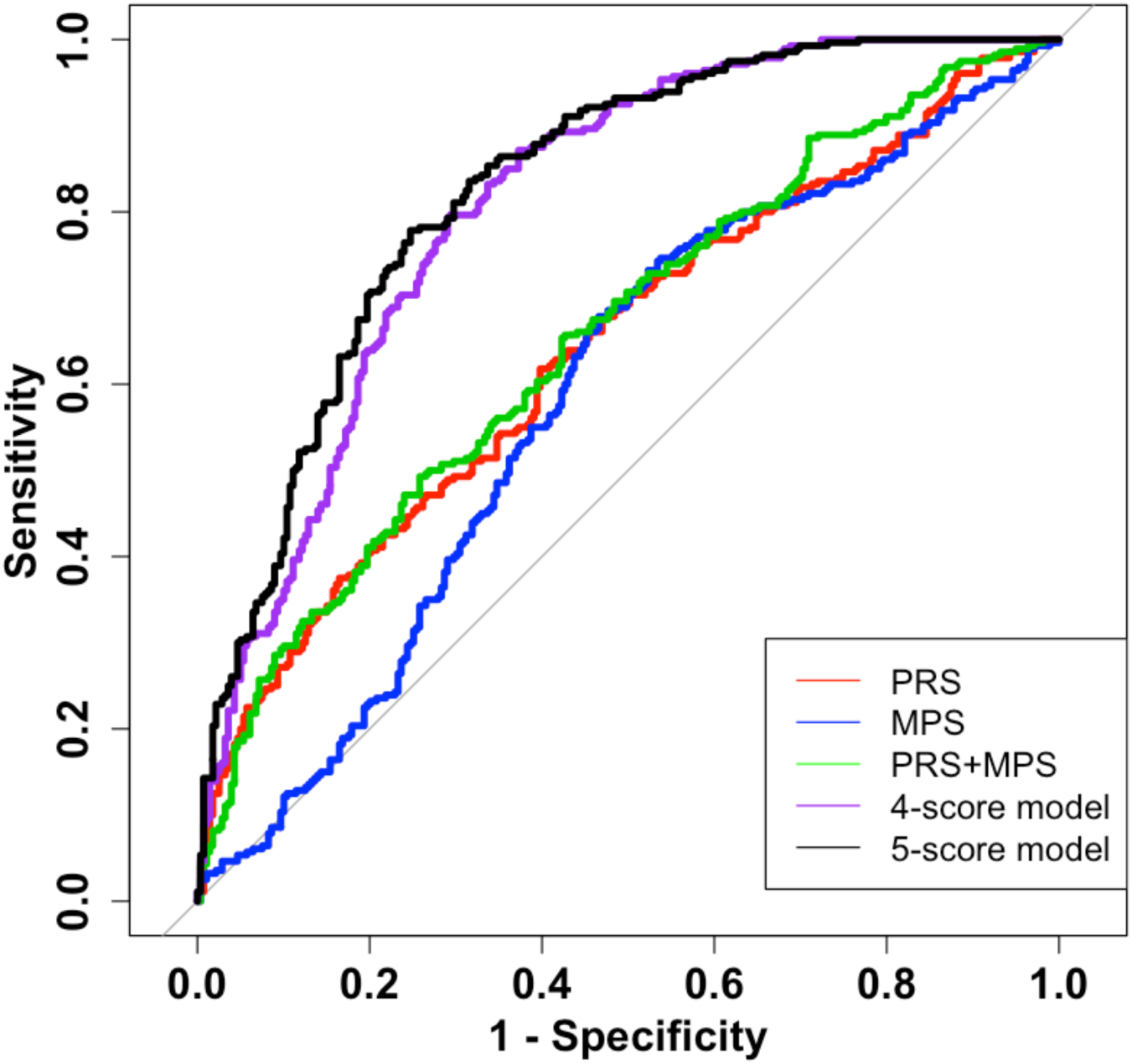
Receiver-operator curves for different multi-score models in the Oslo cohort. The figure illustrates how the performance in differentiating PD from control participants improves as multiple scores are incorporated into the risk profiling model. Lines show one representative iteration out of 1000 where Oslo data were split into Training and Test datasets to estimate model parameters and test out-of-sample classification, respectively. The 4-score model incorporates four different methylation-based scores, namely methylation profile score, smoking score, granulocyte proportion and monocyte proportion. The 5-score model included all these in addition to polygenic risk score. The 5-score model had on average an area under the receiver-operator curve (AUC) of 0.82. (See also Extended Data Table 3). **Abbreviations:** PRS = Polygenic risk score, MPS = Methylation profile score

### Assessing the performance of joint risk profiles in the PPMI baseline data

To evaluate the performance of multi-score profiling to differentiate PD from controls near the time of diagnosis, we calculated equivalent risk profiles in the PPMI dataset and performed logistic regression analyses and out-of-sample classification. Based on the lack of replication for top EWAS probes in PPMI data, we anticipated that methylation-based scores would have less differentiating power in PPMI. In line with these expectations, we generally observed more modest effect sizes than in the Oslo cohort (Table 4). Assessing PRS, MPS, SSc and estimated cell type proportions individually in logistic regression models adjusting for sex and age, the direction of effect was consistent with the Oslo data for all variables except NK cell proportion, but reached nominal significance only for PRS (p=2.8*10^-05^), proportions of granulocytes (p=0.0019), CD4+ T cells (p=0.034), and B-cells (p=0.017) (Table 4). The association with SSc also approached significance in the same direction as the Oslo data (p=0.085). Consistent with the Oslo results, age acceleration based on the epigenetic clock algorithm was not associated with disease status (Table 4).

We assessed the performance in PPMI of a model incorporating all independent scores associated with PD in the Oslo data (PRS, MPS, SSc, granulocyte proportion and monocyte proportion), using the coefficients from the full Oslo analysis as weights. As expected, the score showed a strong association with PD in logistic regression adjusting for age and sex (OR=1.80, 95% CI:1.40 - 2.35, p=6.4*10^-06^), but the power to differentiate patients from control participants was lower than in Oslo data (AUC = 0.65).

## Discussion

We conducted an epigenome-wide association study (EWAS) in whole blood, including 280 participants with PD and 279 healthy controls with European ancestry. We identified 13 differentially methylated probes, but were not able to replicate any of these associations in independent data from baseline visits in the PPMI study. We nominate a hypermethylated CpG in *LPIN1* that passed a FDR threshold in meta-analysis and reached nominal significance in both Oslo, Vallerga et al. and PPMI data as a novel suggestive signal. Furthermore, we found that individual scores for epigenetic PD risk, estimated smoking exposure, leukocyte type proportions and polygenic PD risk were independently associated with PD risk, differentiating patients from controls with an AUC of 0.82 when all significant scores were integrated in a multi-score risk profile. Follow-up in the independent PPMI PD-control cohort sampled at baseline showed a more modest association with a multiscore methylation profile which did not significantly improve out-of-sample classification beyond PRS. Taken together our results shed light on the interpretation of whole blood EWAS and the future biomarker potential of DNA methylation profiling in PD.

We compared our results to the largest whole blood PD EWAS to date, recently published by Vallerga et al (13). Importantly, we assessed differential methylation using the same statistical approach as this study, the MOA method implemented in the OSCA software package, which has been developed to overcome the challenges of potential unobserved confounding and control genomic inflation in EWAS studies (21). However, the Vallerga et al. study used an earlier version of the Illumina methylation array (Human Methylation 450K BeadChip) with fewer probes compared to the latest MethylationEPIC array (19), limiting the opportunities for cross-validation of differentially methylated CpGs.

The top associated probe highlighted by Vallerga et al, cg06690548 at the *SLC7A11* locus (13), was robustly replicated in the Oslo EWAS, but not in PPMI data. Similarly, the coefficients of sub-significant top probes from Vallerga correlate significantly between the Vallerga and Oslo datasets, but not the PPMI results. A plausible interpretation of this discrepancy may be that PPMI patients are sampled at baseline, whereas the other studies included patients after many years of disease duration. PD is a disease with widespread implications on patients’ lifestyle and exposures, including medication, nutritional factors and physical exercise, likely leading to various downstream consequences that affect DNA methylation. These observations illustrate that differential methylation observed in EWAS of complex diseases may reflect effects as well as causes of disease, something that needs to be carefully considered when designing future studies.

In a meta-analysis of MOA results from the Vallerga et al. study, Oslo and PPMI we identified 4 probes passing FDR < 0.05, with cg10142874 at the *LPIN1* locus passing p < 0.05 in all three datasets. A recent study focusing on ferroptosis-related genes identified *LPIN1*, encoding the enzyme Lipin-1, as a candidate PD biomarker applying a machine learning approach to whole blood mRNA expression data, showing significantly reduced plasma levels of *LPIN1* in both early-stage and mid-to late stage PD subjects in an independent validation experiment (37). An increasing body of evidence indicates that *ferroptosis*, a specific type of iron-dependent regulated cell death due to the build-up of harmful lipid peroxides (38), is linked to PD pathogenesis (37, 39, 40). Interestingly, α-synuclein oligomers have been shown to bind to plasma membranes and initiate ferroptosis in human stem cell-derived models of synucleinopathy (40). Our suggestive finding of cg10142874 hypermethylation in PD may be hypothesized to reflect downregulation of *LPIN1*, consistent with the recent machine-learning study (37). Furthermore, hypermethylation of cg06690548 at the *SCL7A11* locus has been associated with decreased levels of the cystine/glutamate antiporter (xCT) (13). A recent review highlighted reduced xCT levels leading to impairment in the antioxidant system as an one of several initiating events in ferroptosis that align with the observed pathological alterations in patients with PD (39), suggesting that *SLC7A11* hypermethylation might also be linked to ferroptosis. More research is warranted to clarify the role of *LPIN1* and *SLC7A11* both in ferroptosis in general and PD pathogenesis specifically.

Epigenome-wide DNA methylation data can currently be used in combination with reference data and software algorithms to estimate scores for both cell type proportions and complex phenotypes such as disease risk, epigenetic aging or past exposure to smoking. We show how several PD-associated scores may be combined in a multiscore model which showed remarkable power to differentiate PD patients from controls in the Oslo data (AUC = 0.82). However, the model was far less powerful in the PPMI data, which may indicate that the profiling captures changes that largely develop after PD diagnosis. Nevertheless, our results provide a compelling proof of principle for the combination of multiple methylation-based scores into a joint risk profile as a promising approach for future studies. We anticipate that the value and power of this methodology may increase also for early or prodromal PD as algorithms improve and large-scale reference data become available for a wider range of phenotypes and exposures.

The methylation scores that associated most strongly with PD status in our analyses were estimated leukocyte proportions. This is in line with many previous studies (13, 18). It may well be argued that conventional leukocyte counts would have captured the same variation in a cheaper and quicker way. However, for research purposes, collecting EDTA blood for DNA extraction is fast and easy to organize. We would expect that for many large-scale studies with centralized biobanks, methylation-based leukocyte proportions may turn out to be a more accessible way to incorporate these variables in future studies aiming to model PD risk.

We observed a negative association between a methylation-based estimate of exposure to tobacco smoke and PD, consistent with epidemiological observations. Others have used a similar scoring approach and included estimated smoking as a covariate in EWAS analysis (13). In contrast, we took advantage of this known PD-associated exposure to show that SSc may also carry potential for improved risk prediction in a model integrating multiple disease-associated scores. This finding provides proof of principle that DNA methylation profiling may be a useful tool to objectively quantify retrospective environmental exposures of relevance to PD risk. With increasing availability of reference data, we anticipate that scoring algorithms could be developed for additional risk factors of interest, such as pesticide exposure, diet or alcohol consumption, which may further improve the potential for methylation-based risk profiling. We also note that the association with SSc may not necessarily be causally driven by smoking alone. Future studies incorporating smoking data may be able to assess whether the SSc captures a methylation pattern that is associated with PD risk even within a group of non-smokers due to shared molecular mechanisms with other risk-modifying processes, thus helping to elucidate the complex causal patterns determining disease susceptibility.

We note that our study has a number of limitations. The PD participants in the Oslo cohort had on average 10 years’ disease duration at the time of blood sampling. Differential methylation present at this stage is probably largely shaped by downstream consequences of the disease, its treatment and associated lifestyle changes. A previous longitudinal study has demonstrated significant methylation changes as PD progresses (41). Consequently, our joint risk profile model based on methylation and polygenic risk scores showed far better discriminatory power as an out of sample classifier in the Oslo cohort than in the PPMI baseline data. Larger studies of de novo and prodromal PD are warranted to identify early methylation changes and further investigate the potential of methylation profiling as a biomarker for early diagnosis. Furthermore, our study did not incorporate data on clinical PD outcomes, which may ultimately be useful for patient stratification and prognostics (42).

Since a wide range of different environmental exposures can affect the methylome over time, we should ideally have included lifestyle and exposure variables in our analysis, as well as more extensive background information about both the PD and control participants’ health. Most larger whole blood EWAS in complex disease to date have been based on available case-control sample sets collected with the aim to study genetics or biochemical biomarkers, and we suspect that unobserved differences in environment may be an important reason for the limited reproducibility of differential methylation signals across studies. A systematic integration of environmental variables in future large-scale studies is therefore highly warranted (43).

We chose to use beta values rather than M values as measure of DNA methylation and tested for association with PD using the MOA method implemented in the OSCA software package. This statistical approach effectively controls for unobserved confounding variables by adjusting for the random effect of total genome-wide DNA methylation, but is not as conservative as the related multi component mixed linear model-based omic association excluding the target (MOMENT) method (21). We chose to align our statistical approach with the largest previous whole blood PD EWAS (13), yet we acknowledge that there is currently no established consensus about the optimal bioinformatic pipeline for data processing and association testing in EWAS.

Our study was conducted in individuals with European ancestry, limiting the transferability and utility of the results to other ethnic groups. Multi-ethnic GWAS and genetic studies of underrepresented populations are currently high on the agenda in the PD field (44, 45). Investigating diverse populations will likely be important also in PD methylation studies, especially when methylation profiling is exploited for the development of prediction models for precision medicine.

We have included sex as a covariate in all analyses. We have not prioritized sex-stratified analyses, which would have had considerably less statistical power. Yet, we note that previous PD methylation studies have argued for female- and male-specific analyses (46) and that this will be warranted when larger datasets become available.

In conclusion, we have performed a whole blood PD EWAS in 559 participants, cross-validated results with the PPMI and Vallerga et al. studies and conducted a meta-analysis based on more than 3000 individuals. Our results indicate that disease stage may be important for the interpretation of differential methylation in PD patients. We observed limited consistency across cohorts, but replicate differential methylation near *SLC7A11* in established PD and nominate cg10142874 in *LPIN1* from the meta-analysis of all three datasets. Furthermore, we explored the potential of methylation-based risk profiling, combining multiple scores in a joint risk model. This approach demonstrates how a single array dataset may be utilized to generate a PD methylation profile score, estimated leukocyte proportions and methylation-based smoking score, all contributing independently to risk modulation. We anticipate increasingly advanced and accurate methylation-based risk modeling as more relevant reference data become available, and a role for DNA methylation data in patient stratification and future precision medicine for PD.

## Methods

### Subjects

The PD and control participants in the Oslo cohort were recruited from 2007-2020 at the Department of Neurology, Oslo University Hospital, Norway (47, 48). All PD patients underwent neurological examination and were diagnosed based on the UK Parkinson’s Disease Society Brain Bank criteria (49), with the exception that more than one affected relative was not regarded an exclusion criterion. Patients have been screened for the *LRRK2* G2019S mutation, and carriers of this variant as well as patients known to have other monogenic forms of PD were excluded. Control subjects were either spouses of patients, outpatients in primary care without neurological disease or healthy volunteers recruited in the community, all without known parkinsonism among first-degree relatives. The Regional Committee for Medical Research Ethics (Oslo, Norway) approved the study, and the local data protection office approved the sample and data collection. Written, informed consent was given by all subjects. Genetic association results from the same study based on sample sets partly overlapping with the present work have been published previously (47, 50). Demographic characteristics are shown in Table 1.

### Methylation analysis and raw data processing

DNA from the Oslo cohort was extracted from whole blood using standard techniques. Following bisulfite conversion of 500ng DNA from each sample, genome-wide DNA methylation was assessed using the Illumina Infinium MethylationEPIC BeadChip (19). Raw data were imported into R 4.3 and processed using the R packages minfi (25) and wateRmelon(51). Sites with a detection p-value > 0.05 were less than 1% in all samples. After filtering out sites with a beadcount < 3 or detection p-value > 0.05 in > 1% of samples, data were converted to a MethylSet including data on 285 controls, 288 PD, and 4 technical replicates across 847,262 probes. We took advantage of sex chromosome probes to estimate sex from methylation data and removed 4 samples that did not match database sex. We further removed 4 samples flagged as outliers by the wateRmelon *outlyx* function and 1 sample with evidence of suboptimal technical quality based on the minfi *getQC* function. Methylation data were normalized using the *dasen* method recommended by the developers of the wateRmelon package (51). After mapping of methylation data to the genome we removed probes on sex chromosomes, probes overlapping loci with known single-nucleotide polymophisms (SNPs) and probes previously reported as cross-reactive (52), leaving a total of 775,795 CpG probes. Previous work has shown that a substantial proportion of CpG probes are unsuited for association testing in complex disorders due to large measurement errors. We took advantage of technical replicates to assess the relative contribution of biological variability to total variability for each probe, computed as the intra-class correlation coefficient (ICC) using the CpGFilter package (53). We removed probes showing predominantly technical variation (ICC < 0.1), leaving 502,261 CpGs in the datset. Finally, constitutively methylated probes with mean beta values > 0.975 or <0.025 and 5 samples lacking age data were filtered out leaving a total of 497,577 probes in 279 controls and 280 PD participants for association testing.

For the PPMI cohort, wholeblood DNA methylation data from two different projects are available in the PPMI online portal (www.ppmi-info.org). We chose to analyze the Illumina MethylationEPIC data from “MJFF project 120”, which included the largest number of PD and control samples from PPMI baseline visits. We intersected IDs from the June 2023 PPMI data cut with the sample list for this methylation project and identified 324 samples with a primary diagnosis of PD and 129 controls. Raw data for these samples were imported into R and processed with an identical QC pipeline as implemented in the analysis of the Oslo cohort. The initial MethylSet dataset included 453 samples and 851076 probes. Three samples were removed due to detection p-value >0.05 in >1% of sites. There were no sex-check failures. The minfi *getQC* function identified one low quality sample. We also excluded samples with known monogenic PD (including *GBA1, LRRK2* and *SNCA*) and other ethnicities than “white”. After filtering out CpGs on sex chromosomes, probes overlapping with SNPs and cross-reactive probes, the final dataset included 779,321 CpGs in 269 PD patients and 109 controls.

### Genotype data

Genotyping, quality control and imputation has been previously described for the Oslo samples (54, 55). In brief, genotyping was performed on the Illumina Infinium OmniExpress24 or NeuroChip array, followed by quality filtering for missingness, excess heterozygosity, ancestry outliers, cryptic relatedness or sex-check failures on the sample level, as well as for genotype rate or Hardy–Weinberg equilibrium departure on the variant level. Genotypes were imputed using the Michigan Imputation Server (56) with default settings and reference data from the Haplotype Reference Consortium (57).

For the PPMI cohort, we used quality-checked whole genome sequencing data accessed through the Accelerating Medicines Partnership Parkinson’s disease (AMP-PD) v.2.5 platform.

### Statistical analyses of differential methylation

The OSCA (omic-data-based complex trait analysis) software (21) was used to investigate differential methylation and perform EWAS. The software has various modules and provides two mixed-linear-model (MLM) omic association analysis methods that can be used to perform EWAS: MLM-based omic association (MOA) and multi-component MLM-based omic association excluding the target (MOMENT). MOA is a reference-free method. In the MLM-based association analysis, MOA accounts for confounding by fitting all probes as random effects and incorporates the correlation between distal probes caused by the confounding (21). MOMENT is an extension of the MOA method that stratifies the probes into multiple random-effect components, and were generated to model a scenario where some probes were more closely associated to the trait examined (21). In an EWAS simulation study, both MOA and MOMENT were more powerful in controlling for false positive rate and family-wise error compared to other methods (21). To compare our findings with results from the latest and largest meta-analysis EWAS on PD (13), we chose to conduct EWAS using MOA and beta-values. We performed fixed-effect meta-analysis using the R package *meta* v5.2-0.

### Generating risk scores from genotypes and methylation data

We used PLINK 2 (58) and the PRSice-2 software (59) to generate polygenic risk scores (PRS), including only SNPs associated with PD at a genome wide significance threshold (p<5*10^-08^). We used independent weights from summary statistics of the Chang et al. meta-analysis (60), based partly on data from 23andMe inc., since the Oslo cohort is incorporated in the largest and most recent PD GWAS meta-analysis (61). Detailed code is available from (www.github.com/pihlstrom). Methylation profile scores (MPS) were generated with the “--score” command in OSCA, using summary statistics from Vallerga et al. as weights (13, 21).

Horvath’s epigenetic clock (26) was used to calculate DNA-methylation (DNAm) age for each participant. A measure of epigenetic age acceleration was generated for each participant following methodology described in Horvath et al. (28), as applied in a recent publication on PD age at onset and epigenetic age acceleration (27). The epigenetic age acceleration measure is calculated by initially performing linear regression of DNAm-age on chronological age in controls and using the subsequent model to calculate predicted DNAm age for all subjects. The following residuals of observed DNAm age minus predicted DNAm age were defined as the epigenetic age acceleration measure (27, 28).

The epigenetic smoking score (SSc) is a continuous DNA-methylation score associated with smoking. It was developed by Elliot et al (31), utilizing weights from 187 CpG sites reported as significantly associated with tobacco smoking in an EWAS by Zeilinger et al.(30). We used the R package EpiSmokEr (32) to calculate the SSc. In our Oslo data, 156 of 187 CpGs to predict the SSc were present for analysis, while 169 out of 187 CpGs were present in the PPMI cohort. Estimated cell type proportions in whole blood were generated with the *minfi* R package, that utilizes Houseman et al.s reference data and algorithm (24, 25).

### Statistical analyses of methylation-based scores

We used R version 4.3.3 to analyze demographic parameters and conduct statistical tests of PRS and methylation-based scores. Two-tailed student’s t-tests were carried out to compare mean cell type proportions and mean smoking scores between PD and control subjects at cohort level. Mann-Whitney U/Wilcoxon Rank sum test was performed to compare smoking score values between never-smoking cases and controls in the Oslo data. Correlation between continuous variables was measured as the Pearson’s correlation coefficient *R*, and significance of the correlation was assessed with a two-tailed t-test.

PRS, MPS, SSc, epigenetic age acceleration and all cell type proportions (granulocytes, monocytes, CD4+ T cells, CD8+ T cells, B-cells, natural killer cells) were tested individually for association with PD case/control status in the Oslo cohort in logistic regression analyses for each variable, controlling for age and sex (Table 1). Next, we assessed the association of each score with PD status in a a joint logistic regression model including PRS, MPS, SSc, granulocytes and monocytes. We excluded the estimated lymphocyte proportions as we found that these contributed substantially to collinearity in the model. Sex and age were included as covariates. This joint model showed no evidence of important collinearity (variance inflation factor 1.01-1.14), nor any signs of ill fit tested with the Hosmer-Lemeshow test for goodness of fit (test statistic = 6.05, degrees of freedom = 8, p=0.64).

To internally validate this model for out-of-sample classification within the Oslo cohort without overfitting, we randomly assigned the participants into “*Training”* and “*Test”* subsets with ratio (0.8/0.2) using the *set.seed* function in R, ensuring proportionality between cases and controls similar to the original dataset. All independent variables of interest (PRS, MPS, SSc, Granulocyte proportion, Monocyte proportion) were controlled for age and sex in the *Training* subset. The intercept and beta coefficients for the variables of interest were then applied to generate joint risk profile scores for each individual in the *Test* dataset. Note that sex and age were included as covariates when determining model coefficients in the *Training* data, but were not used to generate risk profile scores in the *Test* data. However, when reporting the OR for each joint score, we control for age and sex in the *Test* data. We performed 1000 iterations of this process for each model, and report the mean values from these iterations in Extended Data Table 3. To assess the contribution of individual variables to the out-of-sample classification performance, we also made joint models with fewer variables and calculated the AUC (Extended Data Table 3, Figure 4).

To externally validate the joint risk profile models, we used the full Oslo cohort as the training sample to build the model and the independent PPMI cohort as the test sample. The area under the receiver-operator curve (AUC) has been suggested as the best metric for comparison of DNA-methylation profile disease trait scores among various samples (8), and we used this measure for comparison of the scores.

## Data and code availability

Summary statistics from the Oslo cohort are made available at www.github.com/pihlstrom. Raw methylation data will be made publicly available through the Gene Expression Omnibus repository upon publication of the article. The PPMI data are available through an application process from (https://www.ppmi-info.org/access-data-specimens/download-data) (20).

Meta-analysis summary statistics from the Vallerga et al. EWAS are publicly available from (https://cnsgenomics.com/data/vallerga_et_al_2020_nc/) (13). Meta-analysis summary statistics from Chang et al. GWAS, including data from 23andMe Inc., are made available through 23andMe to qualified researchers under an agreement with 23andMe that protects the privacy of the 23andMe participants (https://research.23andme.com/dataset-access/#how-to). All analysis code used to perform this study is made available on Github (www.github.com/pihlstrom).

## Acknowledgements

We wish to express our gratitude to all study participants. I.H.L, M.M.X.T and L.P were funded by grants from the South-Eastern Regional Health Authority, Norway. M.T was founded by the Research Council of Norway. PPMI data used in the preparation of this article were obtained on September 21^st^ 2023 from the Parkinson’s Progression Markers Initiative (PPMI) database (www.ppmi-info.org/access-data-specimens/download-data), RRID:SCR 006431. For up-to-date information on the study, visit www.ppmi-info.org. PPMI – a public-private partnership – is funded by the Michael J. Fox Foundation for Parkinson’s Research and funding partners, including 4D Pharma, Abbvie, AcureX, Allergan, Amathus Therapeutics, Aligning Science Across Parkinson’s, AskBio, Avid Radiopharmaceuticals, BIAL, BioArctic, Biogen, Biohaven, BioLegend, BlueRock Therapeutics, Bristol-Myers Squibb, Calico Labs, Capsida Biotherapeutics, Celgene, Cerevel Therapeutics, Coave Therapeutics, DaCapo Brainscience, Denali, Edmond J. Safra Foundation, Eli Lilly, Gain Therapeutics, GE HealthCare, Genentech, GSK, Golub Capital, Handl Therapeutics, Insitro, Jazz Pharmaceuticals, Johnson & Johnson Innovative Medicine, Lundbeck, Merck, Meso Scale Discovery, Mission Therapeutics, Neurocrine Biosciences, Neuron23, Neuropore, Pfizer, Piramal, Prevail Therapeutics, Roche, Sanofi, Servier, Sun Pharma Advanced Research Company, Takeda, Teva, UCB, Vanqua Bio, Verily, Voyager Therapeutics, the Weston Family Foundation and Yumanity Therapeutics.

The PD GWAS summary statistics used in this study were generated in a meta-analysis including data from 23andMe, Inc. We would like to thank the research participants and employees of 23andMe for making this work possible. The full GWAS summary statistics for the 23andMe discovery data set will be made available through 23andMe to qualified researchers under an agreement with 23andMe that protects the privacy of the 23andMe participants. Please visit https://research.23andme.com/dataset-access/ for more information and to apply to access the data.

Data used in the preparation of this article were obtained from the Accelerating Medicine Partnership® (AMP®) Parkinson’s Disease (AMP PD) Knowledge Platform. For up-to-date information on the study, visit https://www.amp-pd.org. The AMP® PD program is a public-private partnership managed by the Foundation for the National Institutes of Health and funded by the National Institute of Neurological Disorders and Stroke (NINDS) in partnership with the Aligning Science Across Parkinson’s (ASAP) initiative; Celgene Corporation, a subsidiary of Bristol-Myers Squibb Company; GlaxoSmithKline plc (GSK); The Michael J. Fox Foundation for Parkinson’s Research ; Pfizer Inc.; AbbVie Inc.; Sanofi US Services Inc.; and Verily Life Sciences. ACCELERATING MEDICINES PARTNERSHIP and AMP are registered service marks of the U.S. Department of Health and Human Services.

## Extended data

**Extended Data Figure 1.**
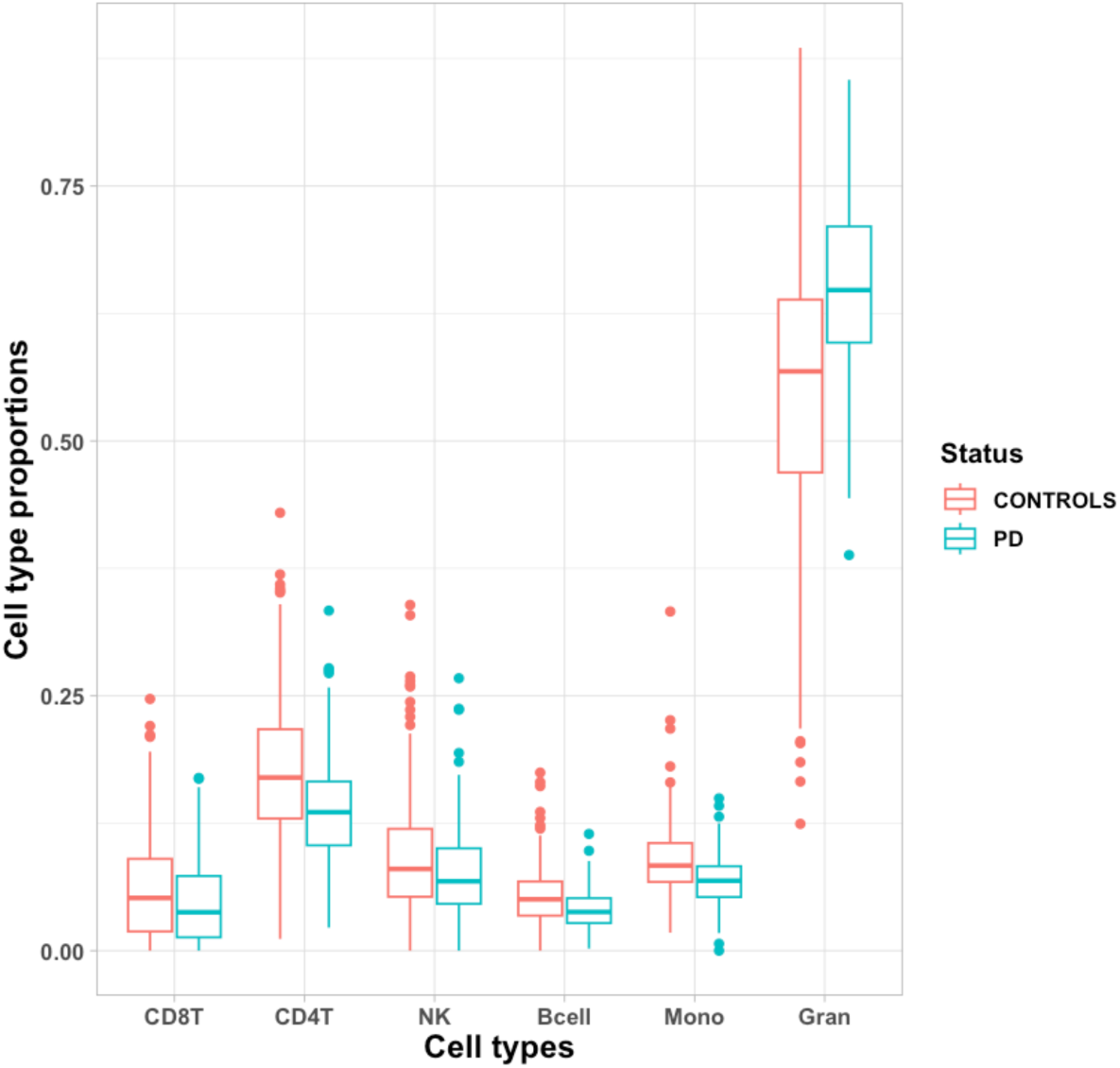
Boxplot of cell type proportions in the Oslo cohort. The boxplot shows cell type proportion estimates for PD participants and controls in the Oslo cohort (N=559). Proportions were estimated from epigenome-wide methylation data using the algorithm from Houseman et al. as implemented in the R package *minfi*. **Abbreviations**: PD = Parkinson’s disease, Gran = Granulocyte proportion, Mono = Monocyte proportion, CD4T = CD4+ T cell proportion, CD8T = CD8+ T cell proportion, Bcell = B-cell proportion, NK = Natural killer cell proportion

**Extended Data Figure 2.**
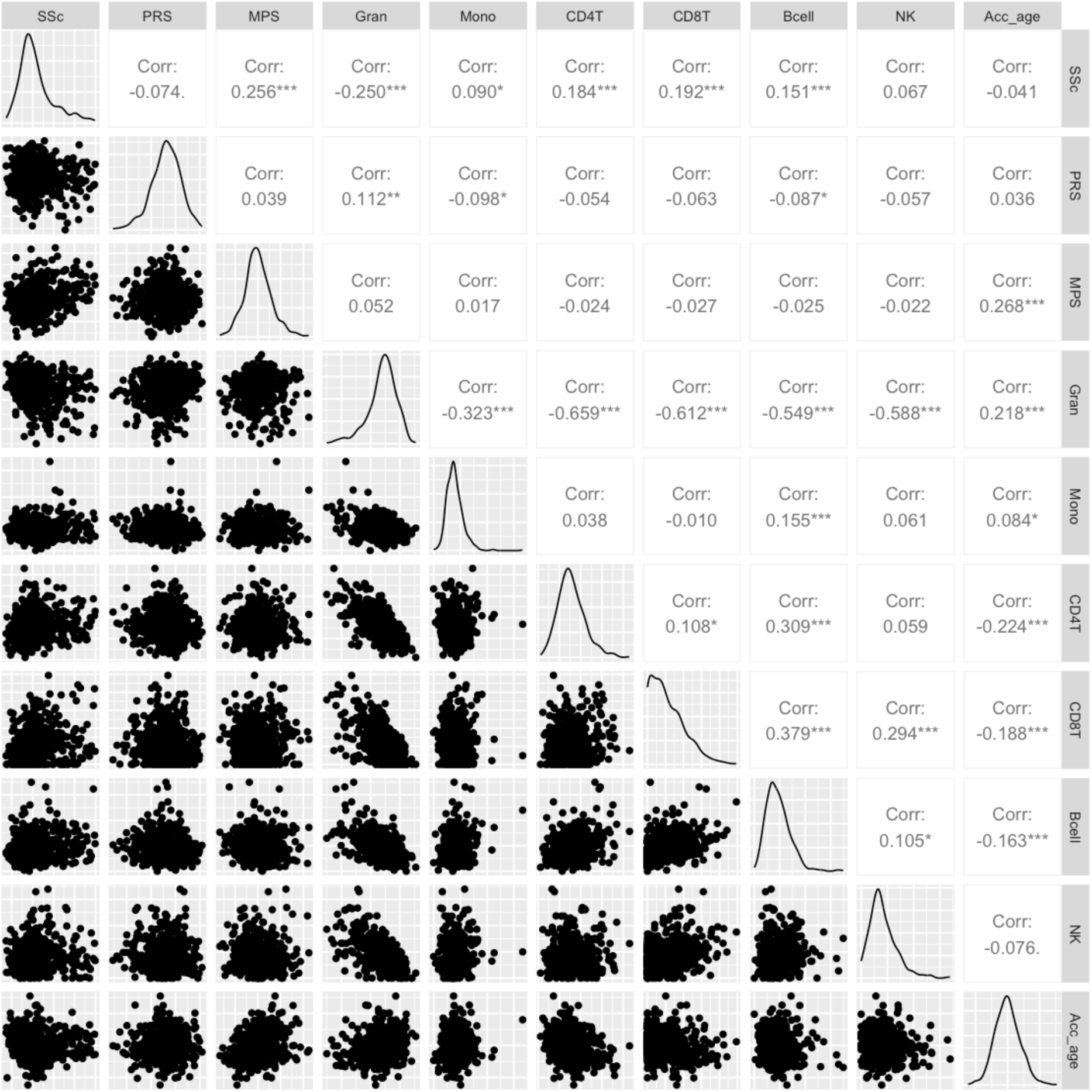
Pairwise correlation (Pearson’s R^2^) of polygenic and methylation-based scores in the Oslo cohort. The figure displays pairwise correlation (Pearson’s R) of polygenic and methylation scores in the Oslo cohort (N=559), in addition to densityplots and scatterplots * = Significant at p <0.05 ** = Significant at p < 0.01 *** = Significant at p < 0.001 **Abbreviations**: SSc = Smoking score (Elliot et al.), PRS = Polygenic risk score, MPS = Methylation profile score, Gran = Granulocyte proportion, Mono = Monocyte proportion, CD4T = CD4+ T cell proportion, CD8T = CD8+ T cell proportion, Bcell = B-cell proportion, NK = Natural killer cell proportion, Acc_age = Accelerated epigenetic age measure

**Extended Data Figure 3.**
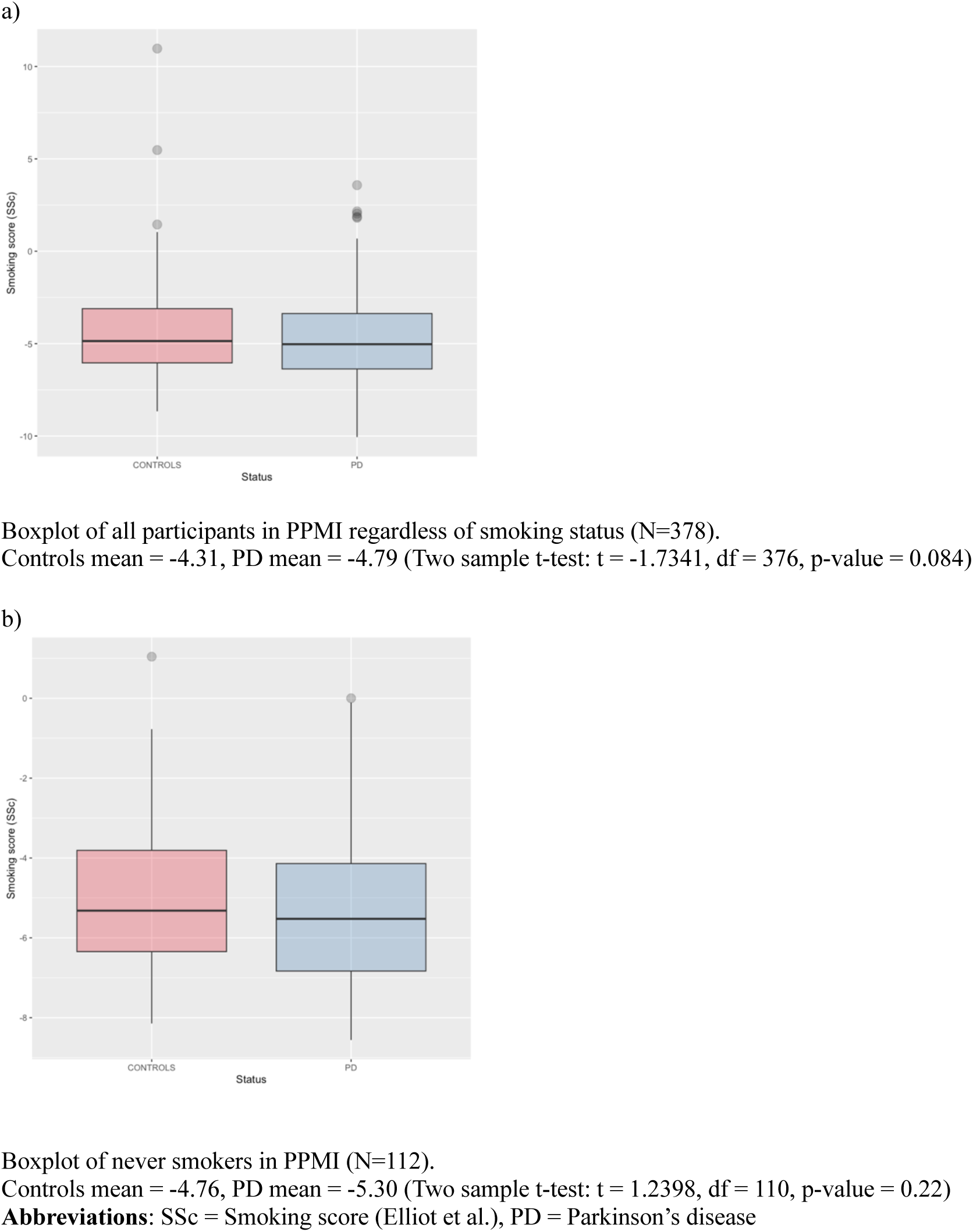
Smoking Score boxplots for the PPMI cohort.

**Extended Data Figure 4.**
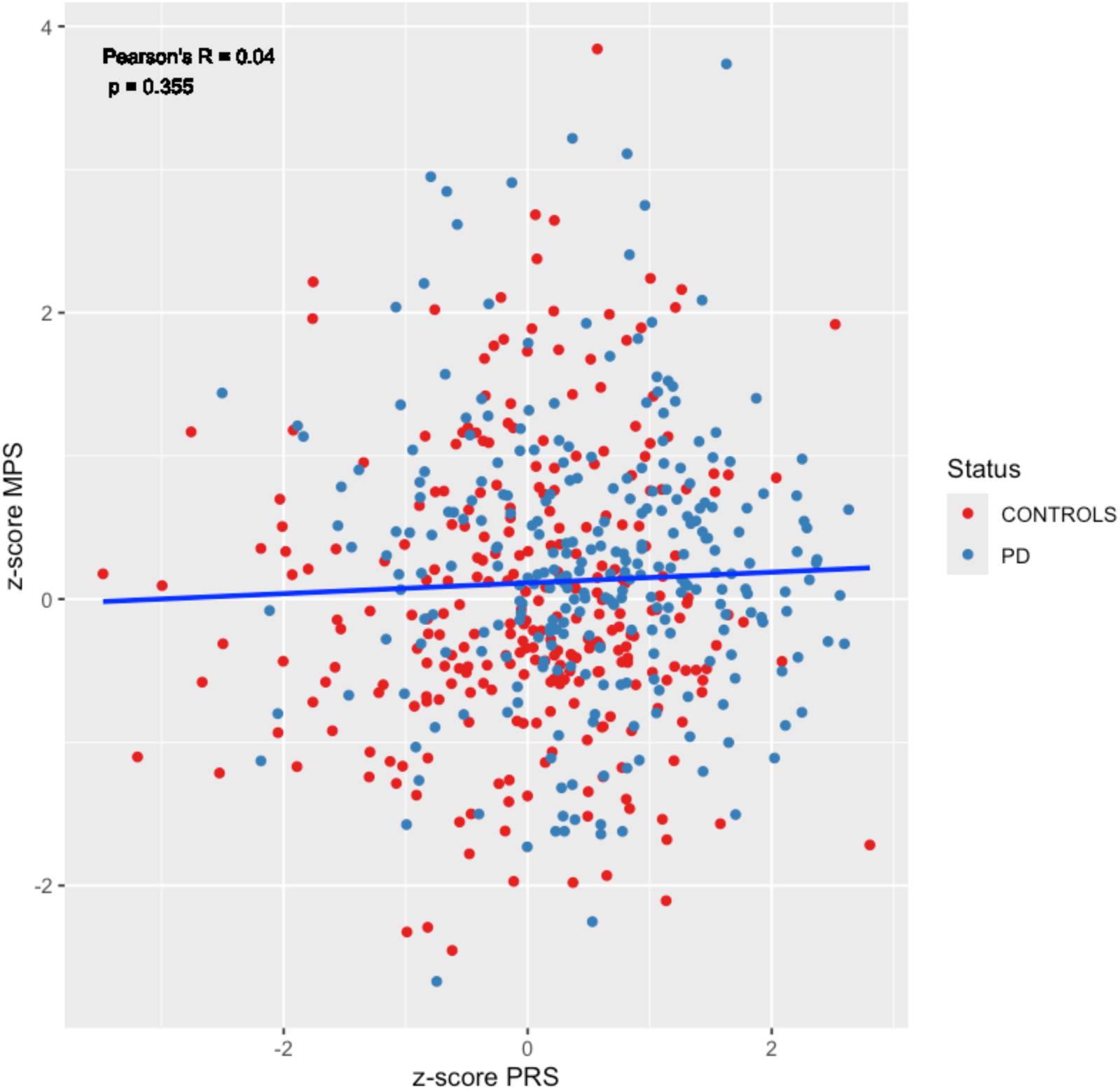
Scatterplot of methylation profile score and polygenic risk score. Scatterplot of methylation profile score and polygenic risk score in the Oslo cohort (N=559). Scores were z-normalized to the mean value in controls. Pearson’s R = 0.04, t = 0.92613, df = 557, p=0.355. **Abbreviations:** z-score MPS = z-normalized methylation profile score, z-score PRS = z-normalized polygenic risk score, PD= Parkinson’s disease

**Extended Data Table 1.**
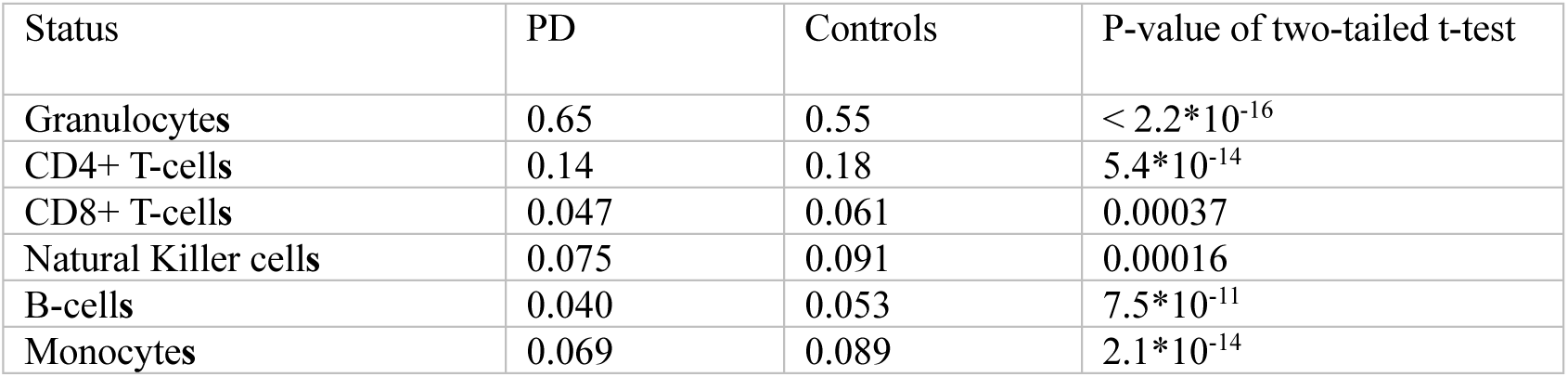
Estimated leukocyte proportions in the Oslo cohort. Leukocyte proportions comparing PD and control participants in the Oslo Cohort (N=559). Blood cell proportions were estimated from epigenome-wide methylation data using the algorithm from Houseman et al. **Abbreviations:** PD = Parkinson’s disease

**Extended Data Table 2.** Logistic regression of a multi-score risk profiling model. Logistic regression was performed in the Oslo cohort data (n=559) with PD status as the outcome variable with all scores fitted jointly in the same model, including sex and age as covariates.

|  | **Estimate**<br>**(B coefficient)** | **Standard**<br>**error** | **z-value** | **p-value** |
| --- | --- | --- | --- | --- |
| Polygenic risk score | 90.48 | 19.05 | 4.75 | $2.0 \times 10^{-06}$ |
| Methylation profile score | 1470 | 390.30 | 3.77 | 0.00017 |
| Smoking score | - 0.16 | 0.03 | -5.30 | $1.2 \times 10^{-07}$ |
| Granulocyte proportion | 7.79 | 1.15 | 6.78 | $1.2 \times 10^{-11}$ |
| Monocyte proportion | - 25.65 | 4.49 | -5.72 | $1.1 \times 10^{-08}$ |

**Extended Data Table 3.**
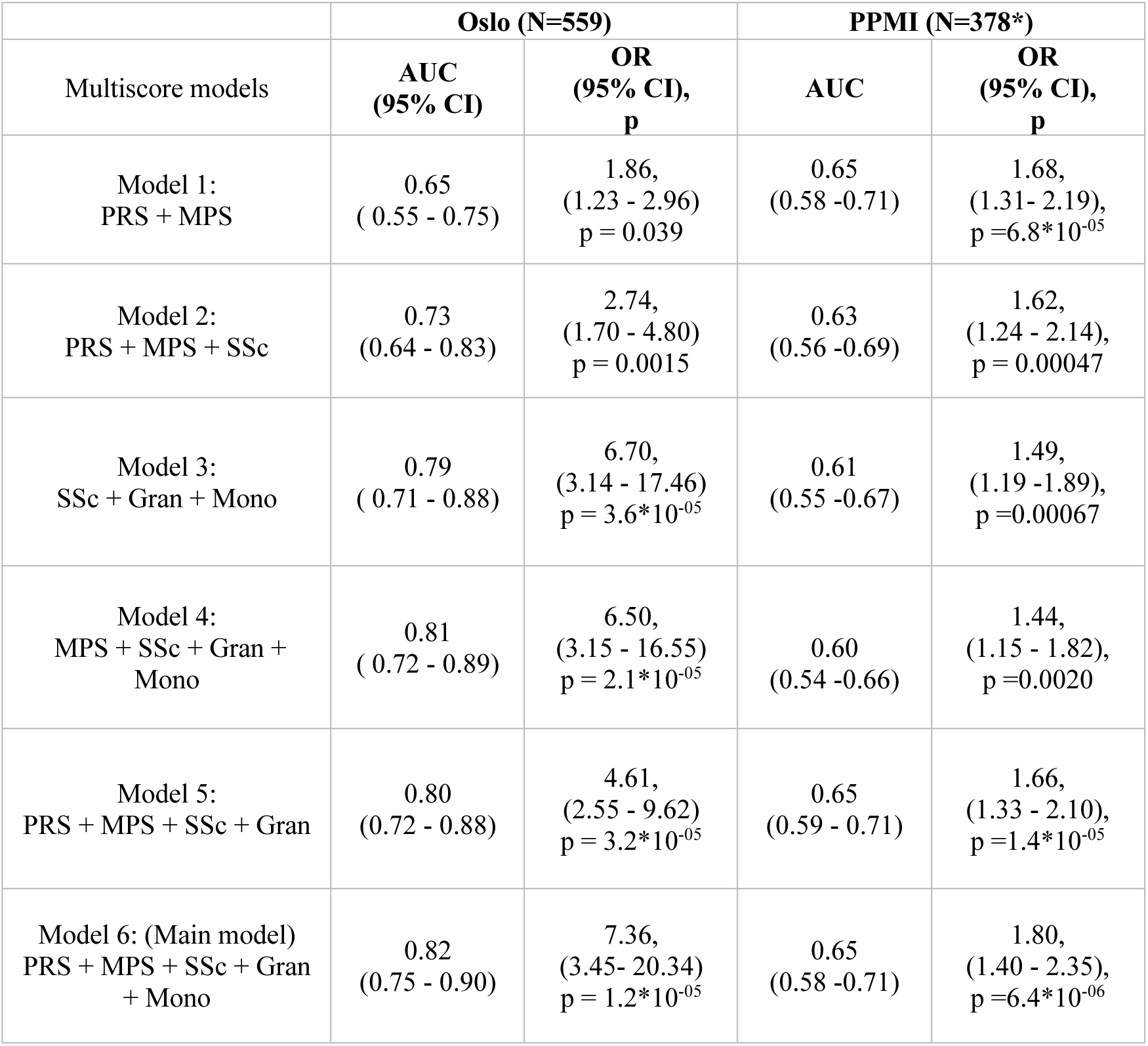
Associations with Parkinson’s disease for joint risk profiles based on different multi-score models. Scores were Z-normalized to the mean value in controls. Odds ratios and P-values are reported for logistic regression models adjusted for age and sex. See main manuscript for further details. **Abbreviations:** P=p-value, AUC= Area under the receiver operator curve, OR = Odds Ratio, 95% CI = 95% Confidence interval, SSc = Smoking score (Elliot et al.), PRS = Polygenic risk score, MPS = Methylation profile score, Gran = Granulocyte proportion, Mono = Monocyte proportion, CD4T = CD4+ T cell proportion, CD8T = CD8+ T cell proportion, Bcell = B-cell proportion, NK = Natural killer cell proportion **\*** In the PPMI-cohort, the logistic regression models including only DNA methylation-scores are performed on n=378 individuals, the models including PRS are performed on N=342 with available genetic data.

## Notes

### Competing Interest Statement

The authors have declared no competing interest.

### Author Declarations

The Regional Committee for Medical and Health Research Ethics, South-East Norway, gave ethical approval for this work.

